# Metabolomic and proteomic signatures of cardiorespiratory fitness for predicting all-cause mortality and non-communicable disease risk: a prospective study in the UK Biobank

**DOI:** 10.1101/2025.08.07.25332939

**Authors:** Sergio Miras-Moreno, Álvaro Torres-Martos, Jonatan R. Ruiz, Jennifer Carter, Carolina Abreu de Carvalho, Concepción M Aguilera, Carmen Piernas, Borja Martinez-Tellez

## Abstract

**Background:** Cardiorespiratory fitness (CRF) is a strong predictor of mortality and non-communicable disease risk, but its underlying molecular mechanisms are poorly understood. In this study, we identified two signatures of CRF (one metabolomic and one proteomic) from UK Biobank participants who completed a risk-stratified submaximal cycle ergometer test, with CRF estimated from the heart rate response to incremental workload.

**Methods:** These signatures were validated in an independent sample of UK participants with data on metabolomics (n=354,222) and proteomics (n=29,961) to investigate prospective associations with all-cause mortality and non-communicable diseases. Prospective associations were evaluated using Cox proportional hazards models adjusted for age, sex, ethnicity, socioeconomic status, lifestyle factors (including smoking, alcohol intake, diet, and BMI), and relevant medical history.

**Results:** Our findings reveal that higher CRF is characterized by downregulation of pathways related to inflammation, triglyceride metabolism, glycolysis, and vascular dysfunction, and upregulation of pathways related to cholesterol transport, apolipoprotein particle size, and cytoskeletal remodeling. Leveraging these insights, we developed two novel signatures of CRF (one metabolomic and one proteomic) that robustly reflect CRF levels (R^2^: 0.50-0.60). Over an average of 9 years of follow-up, we observed 27,659 cases of all-cause mortality. Across the discovery and validation cohorts, we found that the metabolomic signature of CRF was strongly associated with a 39–54% lower risk of all-cause mortality and markedly reduced risk of type 2 diabetes (90% in both), cardiovascular disease (42–47%), and colorectal cancer (33–39%). Additionally, the proteomic signature of CRF was associated with a 17% lower risk of all-cause mortality, and with a 22–39% lower risk of type 2 diabetes and cardiovascular disease.

**Conclusion:** Together, these findings indicate that circulating metabolites and proteins are associated with CRF and with subsequent risk of mortality and non-communicable diseases.

## Introduction

Cardiorespiratory fitness (CRF), defined as the ability of the circulatory and respiratory systems to supply oxygen to skeletal muscles during sustained physical activity, is associated with overall health and functional capacity, and a strong predictor of mortality and non-communicable disease risk^1^. In a landmark study involving over 13,000 participants, Blair et al. were among the first to demonstrate that low CRF was a stronger predictor of all-cause mortality than traditional risk factors such as smoking, hypertension, or high cholesterol^2,3^. These findings have since been replicated and extended, establishing CRF as one of the most powerful modifiable risk factors for premature death^4^. Yet, despite its critical importance, the underlying molecular mechanisms of CRF remain poorly understood. This knowledge gap, coupled with the fact that CRF is rarely assessed in clinical or public health settings, highlights the urgent need for biomarkers to track this vital health indicator.

Given the complex and multifactorial nature of CRF, it is unlikely that a single molecule can serve as an accurate predictor of fitness or related health outcomes. Instead, integrative multi-omic approaches combining large-scale metabolomic and proteomic data are needed to uncover molecular signatures that reflect the physiological adaptations associated with higher CRF. Such signatures may provide novel insights into the biological underpinnings of CRF and offer clinically useful proxies, especially in settings where direct or indirect measurements (e.g., VO₂max testing) are not feasible. Moreover, it is critical to determine whether these molecular signatures are prospectively associated with mortality risk and major health outcomes.

In the present study, we used data from the UK Biobank study^5^ to identify molecular signatures of CRF and subsequently validated these omics signatures for predicting all-cause mortality and non-communicable diseases, including type 2 diabetes, cardiovascular disease, and several types of cancer. We hypothesized that higher CRF is characterized by distinct molecular signatures associated with reduced risk of adverse health outcomes^2,3^. Supported by the unprecedented scale and depth of the UK Biobank, our study provides one of the most detailed omic characterizations of CRF to date. It identifies molecular signatures of CRF with strong predictive value for mortality and non-communicable disease risk and offers new insights into the biological mechanisms and potential clinical applications of CRF.

## Methods

The UK Biobank study was conducted according to the Declaration of Helsinki, and ethical approval was granted by the Northwest Multi-centre Research Ethics Committee (reference number 06/MRE08/65). UK Biobank data available to approved researchers at https://biobank.ndph.ox.ac.uk/showcase/. At recruitment, all participants gave informed consent to participate and be followed up through data linkage. Furthermore, to ensure full transparency and reproducibility, the code used for all analyses is publicly availed in a GitHub repository.

### Study Design and Population

We used data from the UK biobank study, a large-scale prospective cohort that recruited 502,134 participants aged 40-69 years between 2006 and 2010 across the UK (England, Scotland and Wales). At the baseline assessment, participants reported information on sociodemographic factors, medical history and health behaviours through self-completed touchscreen questionnaires. Participants also underwent an examination and a verbal interview with a nurse who collected relevant medical history, blood pressure, height and weight measures as well as blood, saliva and urine samples. In addition, the UK Biobank has established linkages with the NHS Hospital Episode Statistics (HES), cancer registries, and the death registries from the Office of National Statistics (ONS)^5^. The latest UK Biobank protocol is openly available elsewhere (https://www.ukbiobank.ac.uk/wp-content/uploads/2025/01/Main-study-protocol.pdf).

### Metabolomic and proteomic assessment

Metabolomic and proteomic profiling was undertaken in non-fasting blood samples collected at baseline (between 2006-2010). Frozen plasma was extracted from the blood samples collected in EDTA blood collection tubes and used to determine metabolomics using high-throughput nuclear magnetic resonance (NMR) spectroscopy (Nightingale Health Plc, Finland) and proteomic signatures (Olink Analysis Service, Sweden). Metabolomic data is currently available for a subset of around 275,000 UK Biobank participants. Technical details have been previously described ^6^(https://biobank.ndph.ox.ac.uk/showcase/ukb/docs/NMR_companion_phase2.pdf). Targeted metabolomics was used to perform a quantitative analysis, which provided simultaneous quantification of metabolic biomarkers in a single assay expressed in molar concentration units. A total of 251 metabolites were included in the final analyses, including fatty acids and fatty acid composition and lipoproteins in 14 subclasses, amino acids, ketone bodies and glycolysis-related metabolites.

Proteomics data was obtained through the UK Biobank Pharma Proteomics Project, which measured a random sample of 53,058 baseline participants who were representative of the overall UK Biobank population. Information on data processing and quality control can be found in the UK Biobank online resource (https://biobank.ndph.ox.ac.uk/showcase/ukb/docs/Olink_proteomics_data.pdf). Plasma samples were stored at−80 °C and shipped to Olink Analysis Service (Sweden) for analysis. Relative concentrations of 2923 unique proteins were determined using the antibody-based Olink ExploreTM Proximity Extension Assay, in combination with Next-Generation Sequencing. Measurements are expressed as normalized protein expression values (log2-transformed)^7^. All protein levels were standardized in the analyses. After excluding three proteins with a percentage of missing values of >30%, a total of 2920 proteins were included in the proteomic analyses. We excluded 11,680 participants with >10% missing proteomic values as a conservative quality-control criterion to enhance data reliability and reduce the influence of imputation on downstream analyses. The remaining missing values, which were limited in proportion, were singly imputed using a quantile regression approach appropriate for left-censored omics data, implemented with the imputeLCMD package in R. All filtering and imputation procedures were completed prior to model derivation and Cox association analyses.

### Exposure and Outcome definition

The main exposure of interest comprised the metabolomic and proteomic signatures of CRF. The CRF in the UK Biobank was derived via estimated VO₂max from a submaximal cycle ergometer test. Following a pre-exercise cardiovascular evaluation, participants were categorized into four risk groups, which determined the testing protocol: individualized ramp testing for minimal/low risk, constant-load cycling for intermediate risk, and resting ECG assessment for high-risk individuals. Risk stratification was based on a pre-exercise cardiovascular assessment including medical history, current symptoms, medication use, and baseline heart rate and blood pressure measurements. The protocol involved submaximal cycling with continuous heart rate monitoring, either at a constant workload (intermediate risk) or using an individualized ramp protocol (low risk). Heart rate was monitored using a 4-lead ECG device before, during, and after exercise. Estimated maximal workload was obtained by extrapolating the linear heart rate-workload relationship to the age-predicted maximal heart rate (208 − 0.7 × age). The estimated VO₂max was derived using a multivariable equation that incorporated heart rate response, workload, age, sex, and body weight^8^. This non-invasive approach allowed for large-scale CRF assessment in a population-based setting, enabling the inclusion of CRF as a continuous trait in epidemiological analyses. The protocol for measuring fitness is describe in detail in the UK Biobank Cardio Assessment Manual.

The primary outcome of interest was all-cause mortality. Other outcomes of interest included the incidence of fatal and non-fatal cardiovascular disease (CVD), cancer and type 2 diabetes ascertained through linked HES episodes and cancer registries. ICD-10 codes were used to define the outcomes: 1) CVD: coronary heart disease (CHD; I20–I25 and K40-46, K49-50, K75), congestive heart failure or cardiomyopathy (CHF; I50, I50.1, 150.9, I11.0, I13.0, I13.2, I42, I43.1, I43.2, I43.8), and total stroke (I60–I64). Fatal CVD events were measured by I00–I25, I27–I88, and I95–I99; 2) Cancer: breast (C50), colon (C18), colorectal (C18-C20), endometrial (C54.1), bladder (C67), lung (C34) and prostate (C61); 3) type 2 diabetes (E11.0-E11.9), malnutrition-related diabetes (E12.0-E12.9), other specified diabetes (E13.0-E13.9) and unspecified diabetes (E14.0-E14.9).

### Covariate definition

Further details on the derivation and classification of covariates can be found in **Table S1**. Covariates were defined as follows: sex (male, female); ethnicity (grouped as White, Black, Asian, Other); Townsend index of deprivation, a composite measure of deprivation based on unemployment, non-car ownership, non-home ownership and household overcrowding, categorized in quintiles 1-5 (higher quintiles indicating more deprivation); education (grouped as higher degree, any school degree, vocational qualification, unknown); country (England, Scotland, Wales); smoking status (grouped as never, current, previous, unknown); alcohol consumption (grouped as none, occasional, moderate, heavy, unknown); physical activity classified as meeting recommendations (≥150 min of moderate-intensity activity and/or ≥75 min of vigorous activity per week), not meeting recommendations, unknown; dietary quality proxied via meeting fruit and vegetable recommendations (classified as meeting five portions a day, not meeting five portions a day, unknown); body mass index (BMI) categorised as <18.5, 18.5-24.9, 25-29.9, 30-34.9, ≥35 kg/m^2^; family history of CVD, cancer, diabetes (grouped as yes, no, unknown); medication use (grouped as none, lipid-lowering, blood pressure medication); menopause grouped as yes, no, unknown.

### Statistical and bioinformatic analysis

Participant characteristics were studied through descriptive analysis. Categorical variables are reported as total numbers and percentages, whereas continuous variables are reported as means, standard deviations, medians and interquartile ranges.

Retrospective analyses were conducted separately for the metabolomic and proteomic datasets using both univariable and multivariable approaches. Iterative linear models were applied to assess the individual associations between each metabolite and CRF. Nominal p-values were corrected for multiple testing using the false discovery rate (FDR < 0.05) according to the Benjamini–Hochberg procedure. For the metabolomic data, metabolite set enrichment analysis was performed using the fgseaMultilevel function from the fgsea package. Metabolites were ranked according to the beta coefficients derived from the multivariable linear regression model, and enrichment was evaluated across predefined functional metabolite classes based on the UK Biobank metabolomics platform. Normalized enrichment scores (NES) and FDR-adjusted p-values were used to identify significantly enriched metabolic pathways. For the proteomic data, over-representation analysis (ORA) was performed using KEGG pathways on proteins significantly associated with CRF after FDR correction (FDR < 0.05). Protein identifiers were converted to Entrez Gene IDs using the org.Hs.eg.db database, and enriched pathways were identified using the enrichKEGG function from the clusterProfiler package. Statistical significance was assessed using FDR-adjusted p-values. Subsequently, a regularized generalized linear model was used to derive the omics signatures of CRF. This multivariable approach, based on elastic net regularization—which combines L1 and L2 penalties—requires a statistical framework to optimize feature selection and predictive performance. To this end, 10-fold cross-validation was employed to determine the hyperparameters that minimized the root mean squared error (RMSE). The final model was then fitted using the entire dataset and the selected hyperparameters. Calibration of the elastic-net models was further assessed by inspecting residuals versus fitted values plots and by fitting linear models between observed and predicted CRF values to evaluate potential systematic prediction bias (**Fig. S1**). The output from this model was scaled and referred to as the signature of CRF, expressed as z-scores. In both univariable and multivariable analyses, models were adjusted for age, sex, and ethnicity. All analyses were performed in R using the *caret* and *glmnet* packages.

For the prospective analysis of all-cause mortality and health outcomes, we estimated hazard ratios (HR) with 95% CIs for the metabolomic and proteomic signatures of CRF (with scores modelled as quintiles) and the outcomes using multivariable Cox proportional hazard models with age (years) as the underlying time covariate. Person-time of follow-up was calculated from the age at the baseline assessment visit, until the age at which participants were censored, using the date when the outcome occurred. For death, the latest available data was 31 May 2024 for England and Wales; 31 December 2023 for Scotland. For HES-related outcomes, the latest available data was 31 October 2022 for England, 31 May 2022 for Wales, and 31 August 2022 for Scotland. For cancer, the latest available data was 31 December 2020 for England, 31 December 2016 for Wales, and 30 November 2021 for Scotland. Models were sequentially adjusted starting from a minimally adjusted model with age, sex and ethnicity; socio-economic status (Townsend index and education) and country; health behaviours (smoking, alcohol, diet, BMI group); relevant medical history (family history, menopausal status, medication use). The proportional hazards assumption using Schoenfeld residuals’ plots for the exposures was not violated for the variables of interest in the adjusted model (P > 0.05). The survival package in R was used and a two-sided p value of < 0.05 was defined as statistically significant.

## Results

### Participant characteristics

Our initial analyses examined the association between metabolomic and proteomic profiles and CRF in individuals with valid data for either metabolomic or proteomic measures, and CRF. These participants are referred to as the discovery cohort (**Table 1**). The discovery and validation cohorts comprised entirely non-overlapping individuals, ensuring complete independence between datasets. However, partial overlap existed between the metabolomic and proteomic samples due to the larger sample size available for metabolomics within the UK Biobank. The metabolomic discovery cohort included 54,321 individuals (54.9% female) with an average age of 55.9 ± 8.1 years and a mean estimated VO₂max of 28.8 ± 6.7 ml/min/kg. The proteomic discovery cohort comprised 4,235 individuals (54.6% female), with an average age of 55.9 ± 8.2 years and a mean estimated VO₂max of 28.8 ± 6.9 ml/min/kg. After developing the metabolomic and proteomic signatures of CRF in the discovery cohorts, we validated these signatures in an independent group of participants from the UK Biobank—specifically, individuals with valid metabolomic or proteomic data but without available CRF data (named as validation cohort, **Fig. S2**). In this independent cohort, which was followed for an average of 9 years, validation was performed by assessing the association between the predicted metabolomic or proteomic signatures of CRF and all-cause mortality and non-communicable disease risks. During this follow-up period, we documented 27,659 cases of all-cause mortality, 15,267 incident cases of type 2 diabetes, and 40,455 cases of cardiovascular disease. Regarding cancer outcomes, we observed 4,942 cases of colorectal cancer, including 3,341 cases of colon cancer. Additional cancer cases included 9,338 cases of prostate cancer, 8,120 of breast cancer, 3,260 of lung cancer, 1,282 of endometrial cancer, and 906 of bladder cancer. The metabolomic validation cohort included 354,222 participants (56.1% female) with an average age of 55.8 ± 8.1 years, while the proteomic validation cohort comprised 29,961 participants (56.1% female) with an average age of 56.0 ± 8.2 years (see **Table S2**).

**Table 1.**
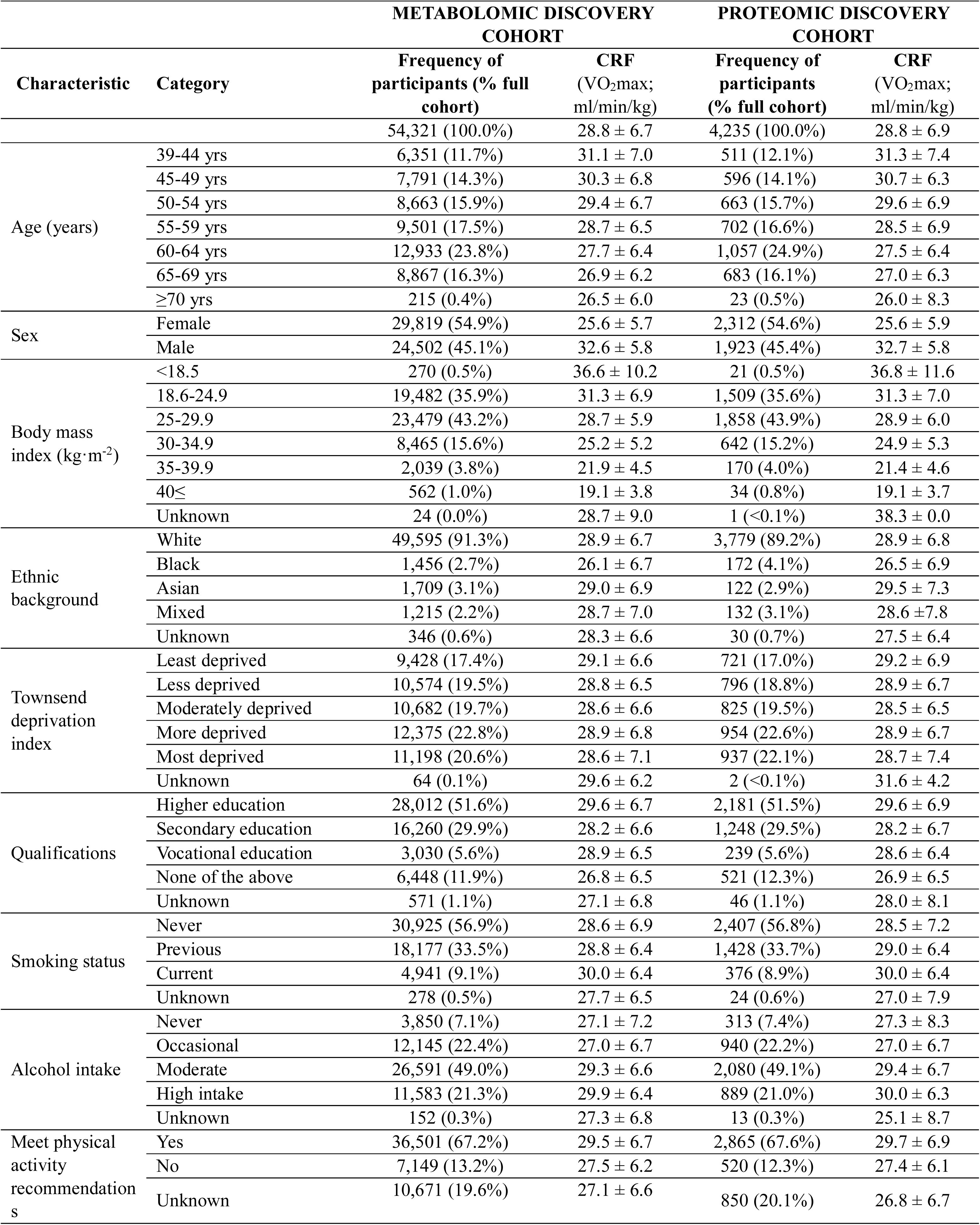
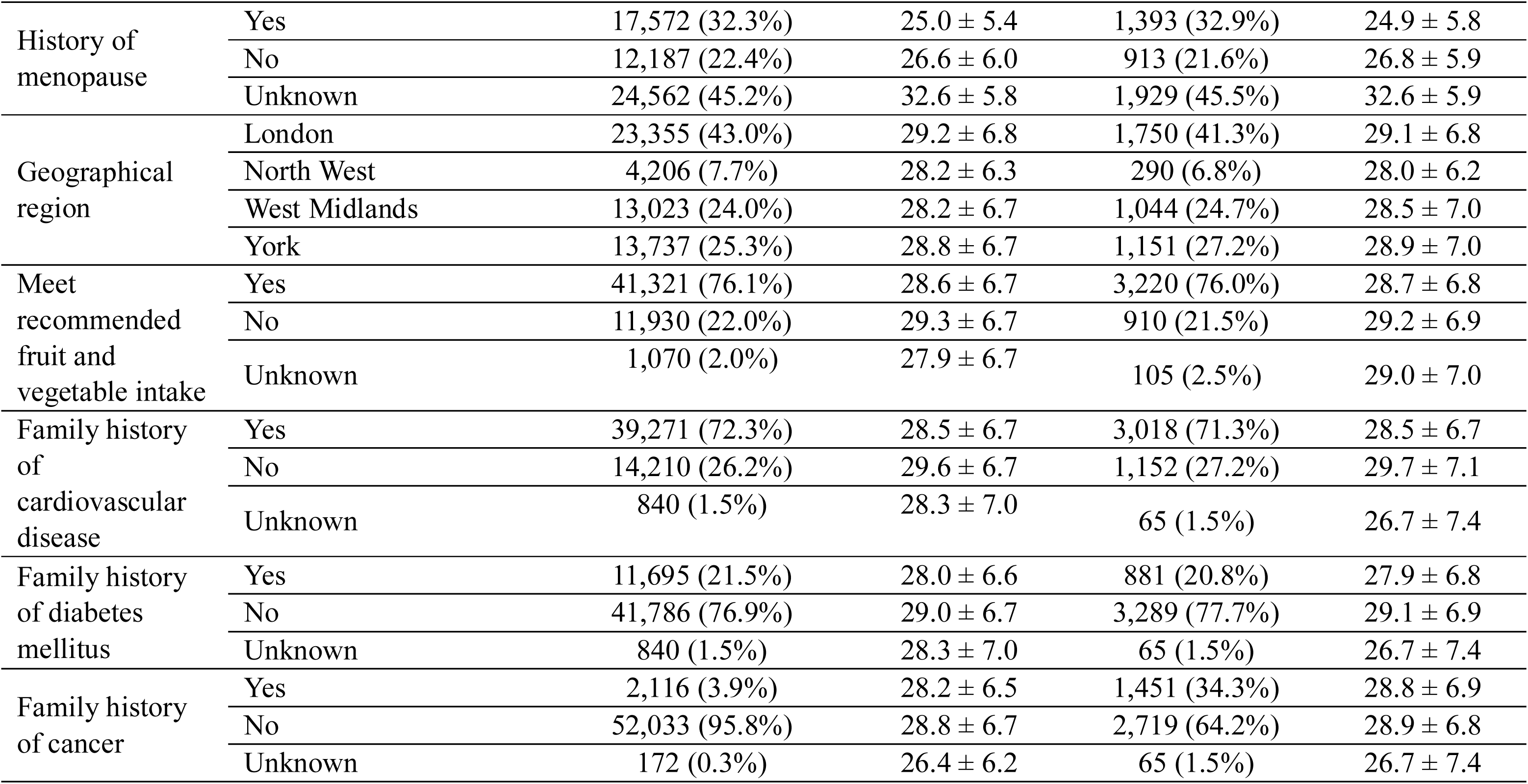
Characteristics of participants included in the metabolomic and proteomic discovery cohorts, and their cardiorespiratory fitness levels (CRF)

### Metabolomic and proteomic profiling identifies widespread molecular associations with cardiorespiratory fitness

In the metabolomic discovery cohort (n = 54,321), we performed multivariable analyses to examine associations between the concentrations of 251 metabolites and CRF levels, adjusting for sex, age, and ethnicity. After False Discovery Rate correction (FDR < 0.05), only 12 metabolites (4.8%) remained non-significantly associated with CRF levels, including acetate, omega-3 fatty acids, and total cholesterol (**Table S3**). However, we found that 239 metabolites (95%) were significantly associated with CRF, of which 156 (65%) showed negative associations and 85 (35%) showed positive associations (**Table S3**). The effect sizes (β) ranged from –0.252 to 0.287. Among the negatively associated metabolites, the percentage of triglycerides within various lipoprotein particles (VLDL, HDL, IDL, and LDL) and VLDL particle sizes were consistently and inversely related to CRF (**Fig. 1A**). Notably, glucose, lactate, pyruvate, and alanine levels were also negatively associated with CRF, though the effect sizes were all β ≤ –0.04 (**Table S3**). Conversely, the percentages of cholesterol and cholesteryl esters within different lipoprotein particles and sizes were consistently and positively associated with CRF (**Fig. 1A** and **Table S3**). We then applied elastic net regression to derive a metabolomic signature of CRF (**Table S4**), retaining 218 metabolites in the model. The 30 most influential metabolites contributing to the metabolomic signature of CRF are shown in **Fig. 1B**, which showed a moderate positive correlation with measured CRF (R² = 0.50; RMSE = 0.71; MAE = 0.53, **Fig. 1C**), supporting the predictive value of the metabolic profile. Using the beta coefficients obtained in **Fig. 1B**, we performed enrichment analyses that revealed that cholesterol was positively associated (NES = +1.56; P = 0.02, **Fig. 1D),** while triglycerides (NES = –1.75; P < 0.01, see **Fig. 1D** and **Table S5**) was negatively associated with CRF.

**Figure 1:**
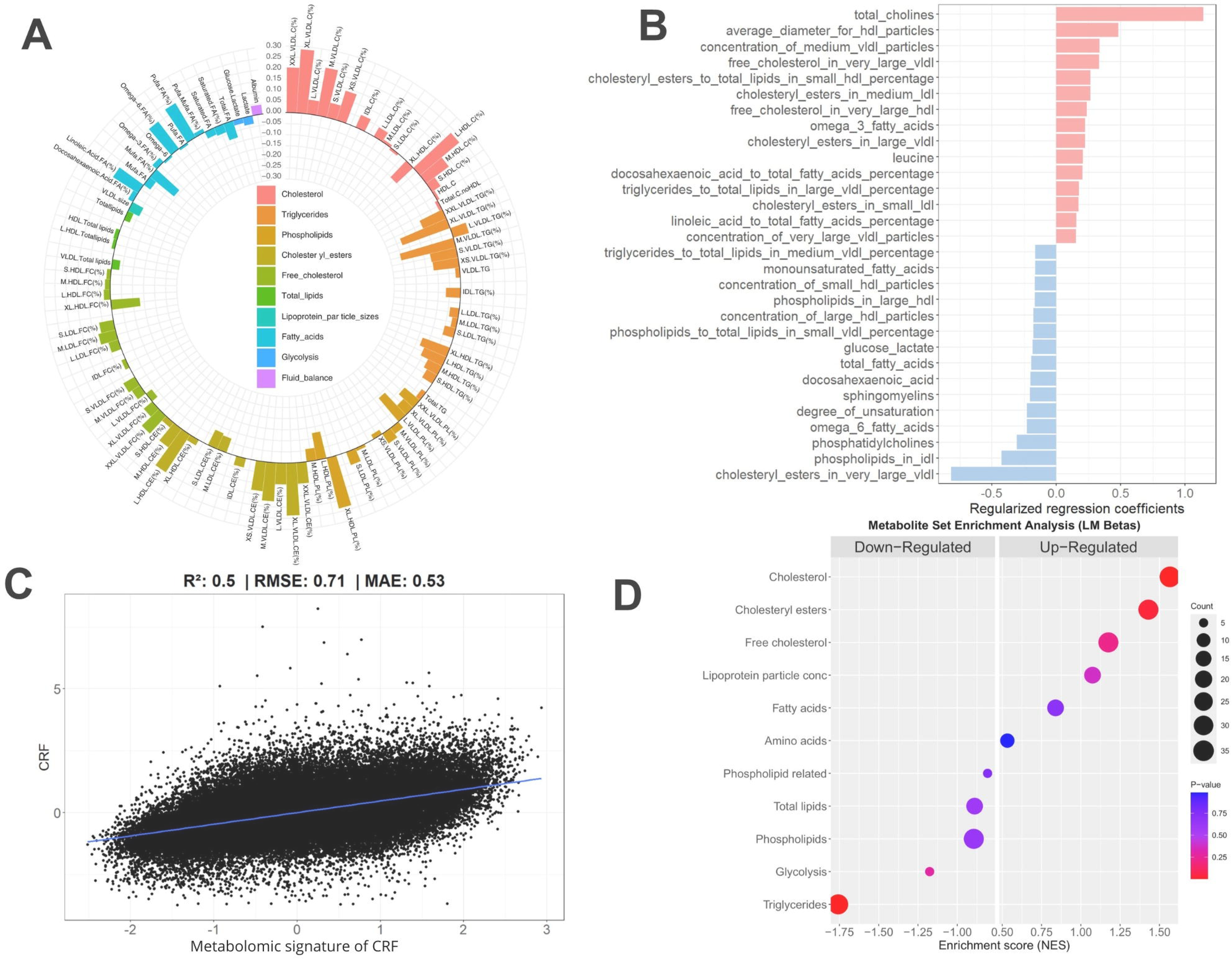
Multivariable associations between metabolomic molecules and cardiorespiratory fitness (CRF) levels in the discovery cohort. (A) Circular bar plot showing individual metabolites significantly associated with CRF (FDR < 0.05) in the discovery cohort. Bars indicate the direction (positive or negative) and magnitude of association. Metabolites are grouped by biochemical class. (B) Top 30 metabolites selected via elastic net regularization as part of the metabolomic signature of CRF. Bars represent the standardized beta coefficients from the final penalized regression model. (C) Scatter plots showing predicted scaled CRF based on metabolomic signatures on the x-axis and scaled CRF on the y-axis. The solid black line represents the line of good agreement between CRF and CRF prediction using metabolomic data. Values indicate the mean predictive performance across 10-fold cross-validation (k = 10). CRF, cardiorespiratory fitness; RMSE, root mean square error; MAE, mean absolute error. (D) Metabolite set enrichment analysis of CRF-associated metabolites ranked according to beta coefficients from the multivariable linear regression models (A). The most significantly enriched metabolic pathways are highlighted. Circle size indicates pathway size, and color denotes FDR-corrected significance. All analyses were performed in the discovery cohort (n = 54,321). Metabolite associations were adjusted for age, sex, and ethnicity. The metabolomic signature of CRF shown in panel B was derived using elastic net models and 10-fold cross-validation to select the hyperparameters that optimized predictive performance. Multiple testing correction was performed using FDR via the Benjamini–Hochberg procedure.

Following a similar approach in the proteomic discovery cohort (n = 4,235), we found that 885 out of 2,922 quantified proteins were not significantly associated with CRF levels (FDR < 0.05). However, 2,037 proteins (69.7%) showed significant associations with CRF, of which 1,715 (58.7%) were negatively associated and 322 (11%) were positively associated (**Fig. 2A** and **Table S6**). Effect sizes were modest, ranging from –0.11 to 0.08. Leptin (LEP) was the protein most strongly and negatively associated with CRF. Proteins negatively associated with CRF showed strong functional enrichment in immune, inflammatory, and vascular pathways (**Table S7**). Over-representation analysis revealed significant enrichment in pathways that were downregulated such as cytokine–cytokine receptor interaction, PI3K-Akt signaling, MAPK signaling, chemokine signaling, and lipid and atherosclerosis (**Fig. 2B**). These were supported by network-based analyses highlighting downregulated proteins involved in platelet activation (e.g., VWF, FGA, VASP), cellular senescence (CDKN1A, FOXO3), and inflammatory signaling (IL1B, MMP9, CCL3) (**Fig. 2C**). In contrast, proteins positively associated with CRF showed weaker enrichment patterns. A smaller set of upregulated pathways emerged, including Ras signaling, regulation of actin cytoskeleton, focal adhesion, and ECM-receptor interaction (**Fig. 2D**). The 30 most influential proteins contributing to the proteomic signature of CRF are shown in **Fig. 2E** and **Table S8**, which showed a strong positive correlation with measured CRF (R² = 0.6; RMSE = 0.62; MAE = 0.46, **Fig. 2F**), supporting the predictive value of the proteomic profile.

**Figure 2:**
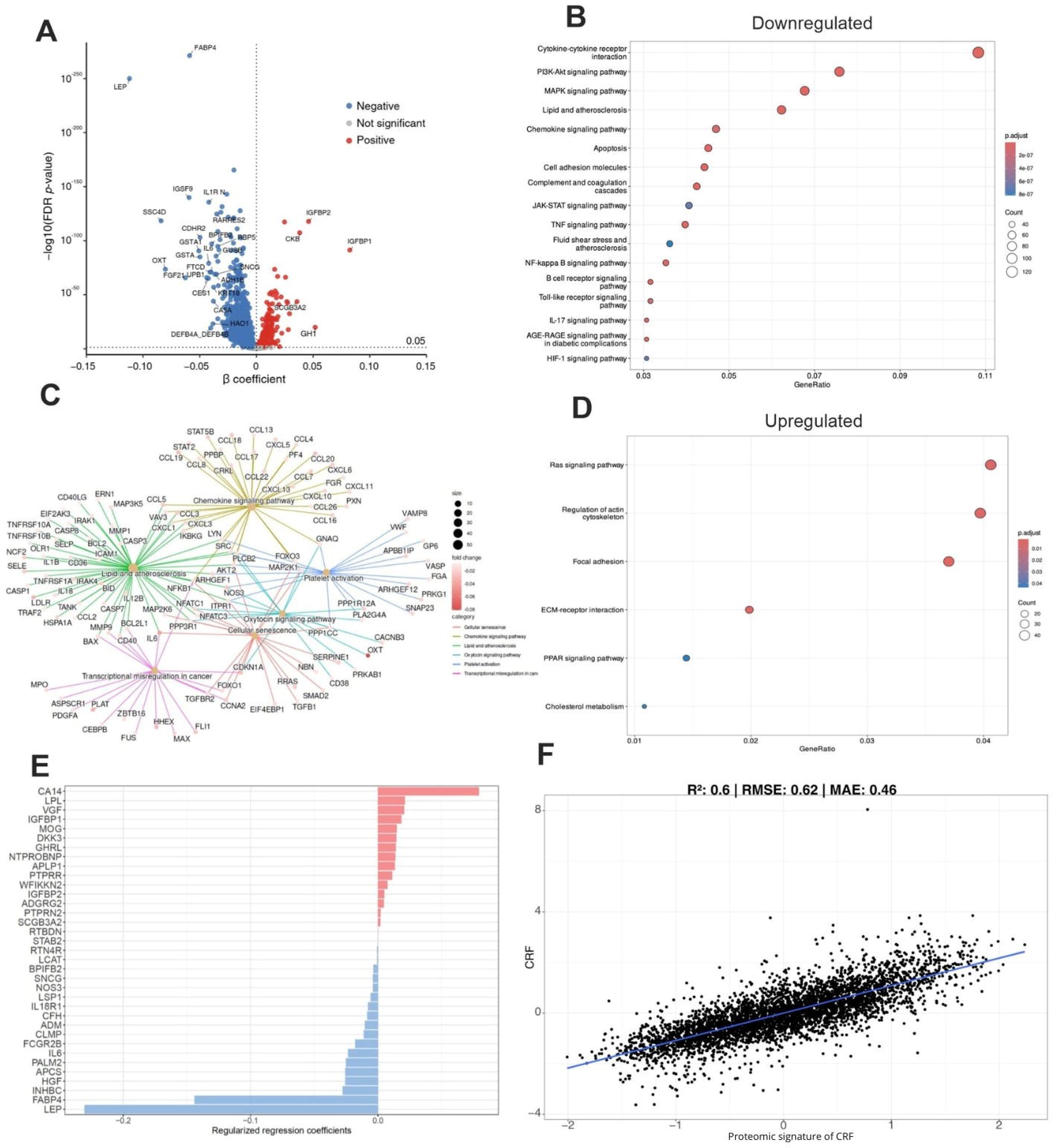
Multivariable associations between proteomics molecules and cardiorespiratory fitness (CRF) levels. (A) Volcano plot displaying the associations between individual plasma proteins and CRF in the discovery cohort. The x-axis shows the β-coefficients from linear models, and the y-axis shows the –log10 FDR-adjusted p-values. Proteins significantly positively associated with CRF are highlighted in red, and those negatively associated are shown in blue (FDR < 0.05). (B and D) Over-representation analysis (ORA) was conducted on proteins significantly associated with CRF. Panel (B) shows enriched KEGG pathways among proteins negatively associated with CRF (downregulated), while panel (D) displays enriched pathways among proteins positively associated with CRF (upregulated). Dot size reflects the number of proteins involved in each pathway (Count), color indicates the adjusted p-value, and the x-axis represents the GeneRatio—defined here as the number of significant proteins in the pathway divided by the total number of proteins analyzed that belong to that pathway. (C) Gene set enrichment analysis (GSEA) based on proteins associated with CRF using standardized coefficients was conducted. Enriched KEGG pathways are represented as colored clusters, and proteins are shown as nodes. Node size indicates the number of proteins of these clusters, while node color intensity reflects the beta coefficient. (E) Top 30 proteins included in the proteomic signature of CRF, selected using elastic net regression with 10-fold cross-validation. Bars represent standardized beta coefficients from the final penalized regression model. (F) Scatter plots showing predicted scaled CRF based on proteomic signatures on the x-axis and scaled CRF on the y-axis. The solid black line represents the line of good agreement between CRF and CRF prediction using proteomic data. Values indicate the mean predictive performance across 10-fold cross-validation (k = 10). CRF, cardiorespiratory fitness; RMSE, root mean square error; MAE, mean absolute error. All analyses were performed in the discovery cohort (n = 4,235). Protein levels were standardized and adjusted for age, sex, and ethnicity. Multiple testing correction was performed using FDR via the Benjamini–Hochberg procedure.

### Metabolomic signature of CRF predicts lower risk of mortality and chronic disease, while the proteomic signature of CRF predicts all-cause mortality, type 2 diabetes and cardiovascular disease

To investigate the association between CRF and the risk of all-cause mortality and non-communicable diseases, we performed the analyses in all participants with available CRF data, regardless of whether metabolomic or proteomic data were available (n = 55,882, see flow chart **Fig. S2**). As expected, CRF was strongly and progressively associated with a lower risk of all-cause mortality. Compared with individuals in the lowest CRF quintile (quintile 1: reference group), those in the highest quintile (quintile 5) had a 22% lower risk of motality (HR = 0.78, 95% CI: 0.66–0.93; P = 0.01; **Fig. 3**). Higher CRF was also associated with lower risk of type 2 diabetes (51%, HR = 0.49, 95% CI: 0.38–0.63; P<0.001 **Fig. 3**), colorectal cancer (54%, HR = 0.46, 95% CI: 0.31–0.67; P<0.001; **Fig. 3**), lung cancer (43%, HR = 0.57, 95% CI: 0.35–0.94; P=0.03 **Table S9**), and colon cancer (59%, HR = 0.41, 95% CI: 0.26–0.65; P<0.001; **Table S9**). No significant associations were observed for cardiovascular disease (**Fig. 3**), prostate cancer, bladder cancer, breast cancer, or endometrial cancer (**Table S9**). In the discovery cohort, the metabolomic signature of CRF showed even stronger associations. Those in the highest quintile had a 64% lower risk of all-cause mortality (HR = 0.46, 95% CI: 0.37–0.57; P<0.001; **Fig. 3**), and markedly lower risk of type 2 diabetes (90%, HR = 0.10, 95% CI: 0.07–0.13; P<0.001; **Fig. 3**), cardiovascular disease (47%, HR = 0.53, 95% CI: 0.42–0.62; P<0.001; **Fig. 3**), and lung cancer (77%, HR = 0.23, 95% CI: 0.13–0.44; P<0.001; **Table S10**). No significant associations were found with colorectal, prostate, bladder, breast, endometrial, or colon cancer (**Table S10**).

**Figure 3:**
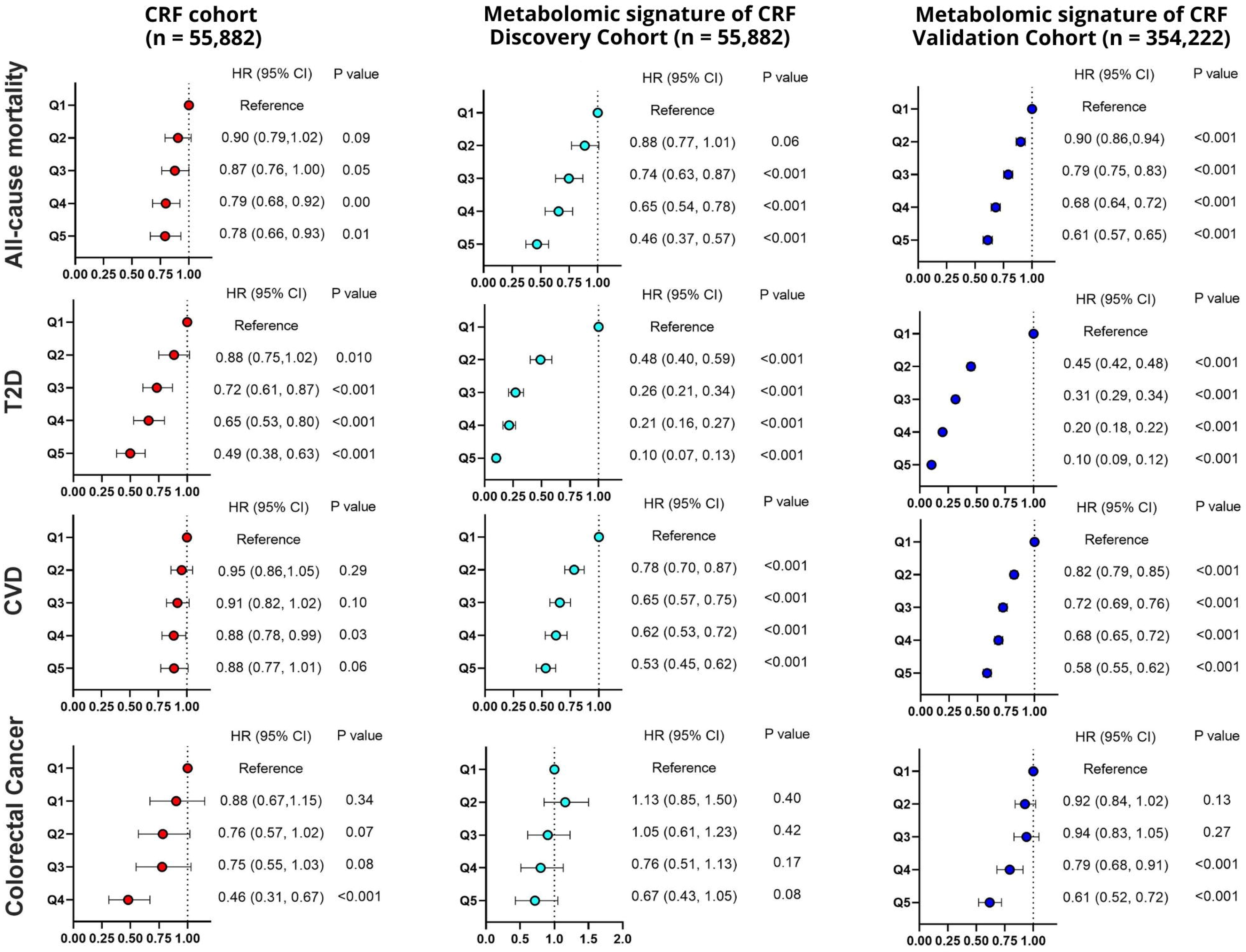
Associations between cardiorespiratory fitness (CRF) and the metabolomic signature of CRF with all-cause mortality, type 2 diabetes (T2D), cardiovascular disease (CVD), and colorectal cancer in the UK biobank population. Hazard ratios (HR) and 95% confidence intervals (CI) are presented across quintiles of estimated CRF and the metabolomic signature of CRF. Results are shown separately for the CRF cohort (left panels), the discovery cohort (middle panels), and the validation cohort (right panel). CRF was categorized into quintiles as follows (n = 55,882): Q1 (n = 11,177), Q2 (n = 11,177), Q3 (n = 11,176), Q4 (n = 11,176), and Q5 (n = 11,176). The number of events (%) for each disease is shown in Table S8. These analyses were performed in all participants with available CRF data, regardless of whether metabolomic or proteomic data were available. For the metabolomic signature of CRF in the discovery cohort (n = 54,321), quintiles were: Q1 (n = 10,865), Q2 (n = 10,864), Q3 (n = 10,864), Q4 (n = 10,864), and Q5 (n = 10,864). The number of events (%) for each disease is shown in Table S9. In the validation cohort (n = 353,908), the metabolomic signature of CRF was similarly divided: Q1 (n = 70,782), Q2 (n = 70,782), Q3 (n = 70,782), Q4 (n = 70,781), and Q5 (n = 70,781). The number of events (%) for each disease is shown in Table S10. All Cox proportional hazards models were adjusted for sex, ethnicity, socio-economic status (Townsend index and education), country, lifestyle behaviours (smoking, alcohol intake, physical activity, fruit and vegetable and saturated fat consumption), BMI category, family history of chronic diseases (diabetes, cardiovascular diseases or cancer), medication use (cholesterol, blood pressure and others) and menopausal status where applicable. Age was used as the underlying time scale. P-values indicate the significance of associations compared to the reference group.

These findings were replicated in the validation cohort (n = 354,322), where the metabolomic signature of CRF was strongly associated with a 39% lower risk of all-cause mortality (HR = 0.61, 95% CI: 0.57–0.65; P<0.001; **Fig. 3**). Participants in the highest quintile had a 90% lower risk of type 2 diabetes (HR = 0.10, 95% CI: 0.09–0.12; P<0.001; **Fig. 3**), as well as significantly lower risk of cardiovascular disease (42%, HR = 0.58, 95% CI: 0.55–0.62; P<0.001; **Fig. 3**), colorectal cancer (39%, HR = 0.61, 95% CI: 0.52–0.72; P<0.001; **Fig. 3**), lung cancer (73%, HR = 0.27, 95% CI: 0.22–0.33; P<0.001; **Table S11**) and colon cancer (46%, HR = 0.54, 95% CI: 0.44–0.66; P<0.001; **Table S11**). No significant associations were observed with bladder, prostate, endometrial, or breast cancer (all Q5 P>0.06; **Table S11**).

The proteomic signature of CRF was associated with a lower risk of type 2 diabetes in the discovery cohort (n = 4,235) (HR = 0.07, 95% CI: 0.02–0.23; P < 0.001; **Fig. 4**). No significant associations were found with all-cause mortality, cardiovascular disease (**Fig. 4**), or any type of cancer (**Table S12**). However, in the validation cohort (n = 29,961), the proteomic signature of CRF was associated with a 17% lower risk of all-cause mortality (HR = 0.83, 95% CI: 0.69–1.00; P = 0.05; Fig. 4), and was significantly associated with a 39% lower risk of type 2 diabetes (HR = 0.61, 95% CI: 0.47–0.80; P < 0.001; **Fig. 4**) and a 22% lower risk of cardiovascular disease (HR = 0.78, 95% CI: 0.66–0.92; P = 0.003; **Fig. 4**). No associations were found with any type of cancer (**Table S13**).

**Figure 4:**
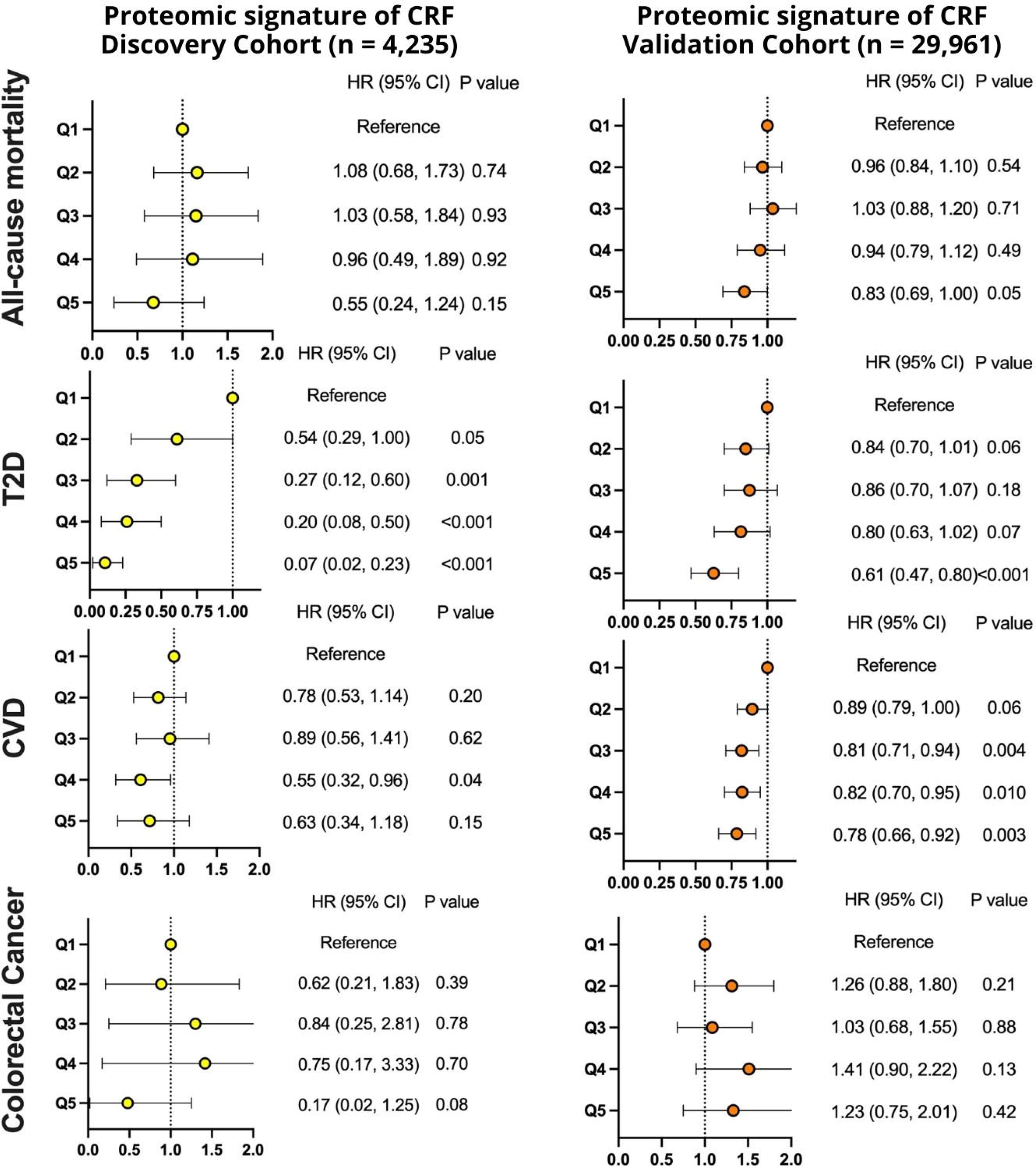
Associations between the proteomic signature of CRF with all-cause mortality, type 2 diabetes (T2D), cardiovascular disease (CVD), and colorectal cancer in the UK biobank population. Hazard ratios (HR) and 95% confidence intervals (CI) are presented across quintiles of the proteomic signature of CRF. Results are shown separately for the discovery cohort (left panels) and the validation cohort (right panel). For the proteomic signature of CRF in the discovery cohort (n = 4,235), quintiles were: Q1 (n = 847), Q2 (n = 847), Q3 (n = 847), Q4 (n = 847), and Q5 (n = 847). The number of events (%) for each disease is shown in Table S11. In the validation cohort (n = 29,961), the proteomic signature of CRF was similarly divided: Q1 (n = 5,993), Q2 (n = 5,992), Q3 (n = 5,992), Q4 (n = 5,992), and Q5 (n = 5,992). The number of events (%) for each disease is shown in Table S12. All Cox proportional hazards models were adjusted for sex, ethnicity, socio-economic status (Townsend index and education), country, lifestyle behaviours (smoking, alcohol intake, physical activity, fruit and vegetable and saturated fat consumption), BMI category, family history of chronic diseases (diabetes, cardiovascular diseases or cancer), medication use (cholesterol, blood pressure and others) and menopausal status where applicable. Age was used as the underlying time scale. P-values indicate the significance of associations compared to the reference group.

## Discussion

In this study, we identified robust metabolomic and proteomic signatures of CRF in a large sample of UK Biobank participants. In short, these signatures reflected that individuals with higher CRF showed downregulation of pathways related to triglyceride metabolism, glycolysis, inflammation, vascular dysfunction, and upregulation of pathways related to cholesterol transport, apolipoprotein particle size, and cytoskeletal remodeling. Both signatures showed good predictive performance, with an R² of 0.50 for the metabolomic signature and R²=0.60 for the proteomic signature. Notably, the metabolomic signature of CRF was strongly and consistently associated with lower risk of all-cause mortality and multiple non-communicable diseases, including type 2 diabetes, cardiovascular disease, and several cancers, whereas the proteomic signature of CRF predicted all-cause mortality, type 2 diabetes and cardiovascular disease. Together, these findings suggest that circulating metabolites and proteins capture the physiological imprint of CRF and may serve as indirect biomarkers for predicting disease and mortality risk.

### Cardiorespiratory fitness is inversely associated with adverse metabolic and inflammatory signatures and positively linked to mitochondrial function

In this study, we investigated the relationship between a broad panel of metabolites and proteins and CRF in 54,321 and 4,235 participants from the UK Biobank, respectively. Specifically, higher CRF was strongly associated with greater fat oxidation capacity and increased activity of the TCA cycle. It is well established that exercise modifies lipoprotein composition and size^9,10^. For instance, a single session of aerobic exercise reduces the triglyceride content within lipoprotein particles, suggesting that triglycerides are being used as an energy substrate during exercise^11^. Similarly, long-term exercise interventions have been shown to reduce atherogenic lipoproteins such as LDL, VLDL, and IDL, as well as other circulating lipid species^12^, highlighting a potential mechanism through which exercise lowers cardiovascular risk and contributes to longevity^10^. Our findings, from the largest study to date combining metabolomics and CRF, reinforce these observations. These results are consistent with previous studies showing that increased mitochondrial density and function are linked to improved cardiometabolic outcomes^13^. Mitochondria are the central organelles for energy production, hosting key metabolic processes such as β-oxidation, the TCA cycle, and lactate oxidation^14^. Furthermore, the proteomic signature of CRF revealed downregulation of inflammatory, vascular, and senescence-related pathways, providing molecular evidence that higher CRF may counteract systemic inflammation and vascular aging. Taken together, these findings support the hypothesis that aerobic (endurance) exercise, through its well-documented ability to increase CRF, may extend lifespan by promoting profound metabolic adaptations. This idea is supported by compelling epidemiological evidence: a robust Finnish cohort study of elite athletes active between 1920 and 1965 showed that endurance athletes lived longer than both power athletes and controls, with significantly lower standardized mortality ratios from all causes and cardiovascular disease (43% and 44% lower, respectively)^15^. Further analyses revealed that these former elite endurance athletes also had a substantially reduced need for chronic disease medications, including a 30–60% lower probability of using drugs for hypertension and type 2 diabetes^16^. The molecular mechanisms underlying these associations are well characterized: aerobic exercise activates AMP-activated protein kinase (AMPK) signaling in skeletal muscle, leading to increased mitochondrial biogenesis^17^. This promotes more efficient utilization of fat and glucose as energy substrates, contributing to better health span and quality of life—and potentially explaining the increased longevity observed in endurance-trained individuals. This notion is further supported by the metabolomic and proteomic signatures of CRF identified in the present study, which reflect enhanced mitochondrial function and substrate utilization.

It is noteworthy that our proteomic signature of CRF highlights similar biological pathways to those observed in the metabolomic signature of CRF. Specifically, individuals with higher CRF showed enrichment in pathways related to energy metabolism, oxidative stress, inflammatory signaling, and cellular regulation, as well as mitochondrial function, angiogenesis, and immune homeostasis. Notably, several of these pathways were also reported in a previous study that identified a distinct proteomic signature of CRF^18^. Interestingly, we observed overlap in both pathways and individual proteins, with six proteins, LEP, FABP4, ADM, TIMP4, FCGR2B, and ANGPTL2, consistently highlighted in both studies.

### Metabolomic and proteomic signatures of CRF as markers of mortality and non-communicable diseases

One of the most striking findings of our study is the strong and consistent association between the metabolomic signature of CRF and reduced risk of all-cause mortality and multiple non-communicable diseases, including type 2 diabetes, cardiovascular disease, and several cancers. In both the discovery and validation cohorts, comprising over 400,000 participants, individuals in the highest quintile of the metabolomic signature of CRF exhibited substantial reductions in mortality (39-54%), cardiovascular disease (42–47%), colorectal cancer (33-39%) and lung cancer (73%). Most notably, they showed up to a 90% lower risk of developing type 2 diabetes. This effect size exceeds those reported in previous studies, where individual metabolomic predictors, such as branched-chain and aromatic amino acids, lipids, and acylcarnitines, have been associated with type 2 diabetes risk, typically with HR ranging from +44% to +400%, depending on the biomarker and cohort. For instance, the Framingham Offspring Study reported up to a fivefold increase in type 2 diabetes risk among individuals with elevated amino acid levels^19^, whereas the EPIC-Potsdam cohort observed a 69% risk reduction in participants with favourable metabolic profiles^20^. Surprisingly, in the UK Biobank cohort, CRF levels were borderline associated with cardiovascular disease risk^8^. Detailed analyses further showed no significant associations with stroke, ischemic heart disease, or atrial fibrillation^8^. In contrast, our metabolomic signature of CRF, derived from 218 circulating metabolites, likely reflects the integration of multiple protective metabolic pathways and appears to outperform previously reported biomarkers. These findings underscore the potential utility of the metabolomic signature of CRF as a biomarker of physiological fitness, capable of capturing molecular features that reflect both metabolic efficiency and long-term disease risk. Notably, this signature showed stronger and more consistent associations across multiple outcomes than traditional CRF measures, suggesting added predictive value.

Despite the strong correlation between measured CRF and the proteomic signature of CRF, we found that the proteomic signature was associated with a lower risk of all-cause mortality, type 2 diabetes and cardiovascular disease, but not with any type of cancer. This is consistent with our proteomic analyses, which showed that IGFBP2 and IGFBP1^21,22^, two proteins positively associated with CRF, are known to enhance insulin sensitivity. These findings align with those of a previous study with a larger discovery sample (n = 14,000)^18^, compared to our cohort of 4,000 participants. Importantly, both the metabolomic and proteomic signatures of CRF should be validated in other cohorts as well as in intervention studies where CRF is modified through exercise training, to determine whether these molecular profiles are responsive to changes in fitness.

Several limitations should be considered when interpreting our findings. First, CRF was estimated using submaximal exercise tests, which rely on extrapolating VO₂max based on heart rate responses. This approach, while practical for large-scale cohorts such as the UK Biobank, may introduce variability due to individual differences in heart rate responses to exercise and factors like medication use, stress, or hydration status. Second, different CRF assessment protocols were applied across UK Biobank subgroups (e.g., varying ramp rates and cycle ergometer loads), which could introduce measurement heterogeneity and limit comparability. Third, not all participants with CRF measurements had available metabolomic or proteomic data. These omics datasets are being generated progressively, and current analyses are limited to a subset of participants, particularly for proteomics, where only ∼4,000 individuals were included in the discovery cohort, potentially limiting statistical power and generalizability. Fourth, outcomes in the UK Biobank were ascertained through national health record linkage. While this method is objective and minimizes selective loss to follow-up, it is susceptible to delayed case detection, often capturing only advanced or severe cases. This may introduce bias toward disease severity and lead to underestimation of associations for milder or earlier disease stages. Fifth, a key limitation of this study is the limited ethnic diversity of the UK Biobank cohort, which comprises predominantly White participants (>90%). Therefore, the generalizability of these findings to more diverse populations may be limited, and replication in multi-ethnic cohorts is warranted. Sixth, metabolomic and proteomic data were derived from non-fasting samples collected at a single time point, which may introduce dietary-related variability and precludes assessment of longitudinal changes or causal inference. Complementarily, it should be noted that elastic-net models were intentionally not adjusted for medication use in order to preserve biological specificity and reduce bias, whereas medication use was subsequently included in the survival models^23^. An additional limitation is the modest discriminatory performance of both omics-derived scores, particularly the weaker performance of the proteomic score compared with the metabolomic score. This fact may reflect the smaller sample size available for proteomic model derivation and the inherent potential error introduced by estimating CRF from heart rate. Despite these limitations, the large sample size, rigorous statistical modeling, and replication of findings in independent cohorts strengthen the validity and robustness of our conclusions

## Conclusions

We identified novel metabolomic and proteomic signatures strongly associated with CRF levels. The metabolomic signature of CRF emerged as a robust predictor of all-cause mortality and multiple non-communicable diseases, including type 2 diabetes, cardiovascular disease, and several cancers. In addition, the proteomic signature of CRF was associated with all-cause mortality, type 2 diabetes and cardiovascular disease. Together, these findings indicate that circulating metabolites and proteins are associated with CRF and with subsequent risk of mortality and non-communicable diseases.

## Data Availability

All data produced are available online at UK biobank

## Acknowledgments

The authors acknowledge the UK Biobank participants and personnel for their contributions to the data resource used in this study (application number 124330).

## Supplemental Materials List

Supplemental Methods:

Table S1

Fig. S1

Supplemental Results:

Tables S2-S13

Fig. S2

## Non-standard Abbreviations and Acronyms

CRF: Cardiorespiratory fitness
HES: NHS Hospital Episode Statistics
ONS: Office of National Statistics
NES: Normalized Enrichment Scores
ORA: Over-Representation Analysis
RMSE: Root Mean Squared Error
MAE: Mean Absolute Error

